# Genetic evidence for repurposing immunomodulatory drugs for major depressive disorder

**DOI:** 10.64898/2026.02.07.26345798

**Authors:** Jehanita Jesuthasan, Abigail R. ter Kuile, Jonathan Roiser, Livia A. Carvalho, Alexandra Pitman, Sandesh Chopade, Chris Finan, Amand Floriaan Schmidt, Jean-Baptiste Pingault

**Affiliations:** Institute of Cognitive Neuroscience, University College London, UK; Department of Clinical, Educational and Health Psychology, University College London, UK; Translational Pharmacology Lab, Department of Clinical Pharmacology and Precision Medicine, William Harvey Research Institute, Queen Mary University of London, Charterhouse Square, London, UK; Division of Psychiatry, University College London, UK; North London NHS Foundation Trust, London, UK; Institute of Cardiovascular Science, Faculty of Population Health Sciences, University College London, London, UK; UCL British Heart Foundation Centre of Excellence, 69-75 Chenies Mews, London WC1E 6HX, United Kingdom; The National Institute for Health Research (NIHR) University College London Hospitals (UCLH) Biomedical Research Centre (BRC), University College London, London, UK; Department of Cardiology, Amsterdam Cardiovascular Science, Amsterdam University Medical Centres, University of Amsterdam, Amsterdam, The Netherlands; Social Genetic and Developmental Psychiatry, King’s College London, London, United Kingdom

## Abstract

**Objective:** To identify immunomodulatory drug targets with genetic evidence in major depressive disorder (MDD), probe symptom-level heterogeneity in their effects, and identify drug repurposing opportunities.

**Methods:** We used *cis*-Mendelian randomisation to evaluate the targets of 204 immunomodulatory compounds, including immunosuppressants, cytokine inhibitors, and anti-infectives. As exposures, we selected genetic instruments from nine genome-wide association studies (GWASs) of protein or gene transcript levels in blood and six brain regions, capturing peripheral and central inflammatory processes. As outcomes, we used summary statistics from GWASs of MDD and eight individual depressive symptoms. We prioritised targets based on consistency across tissues and robustness of Mendelian randomisation estimates, and prioritised compounds based on predicted therapeutic effects.

**Results:** We prioritised 13 drug targets (C1S, CRBN, CUL4A, DEPE1, FCGRT, FKBP1A, HRH1, IL1RL2, IMPDH1, MMP7, POLE2, PRIM1, and S1PR4) associated with MDD risk. These are targeted by 46 compounds – mostly inhibitory – including doxycycline, sutimlimab, and pomalidomide, which may have therapeutic benefits for MDD. In symptom-level analyses, most targets showed heterogeneity across symptoms. Among prioritised targets, three showed consistent effects across more than half the symptoms, while the remainder showed symptom-specific or opposing effects between symptoms.

**Conclusion:** We provide genetic evidence supporting the repurposing of immunomodulatory drugs for MDD, including compounds acting on novel therapeutic targets. Effects differ across depressive symptoms, suggesting that symptoms respond to different drug mechanisms. These findings highlight the importance of considering individual symptoms, rather than MDD as a unitary condition, when developing treatments with a broad spectrum of action or targeting specific symptom profiles.

## Introduction

Major Depressive Disorder (MDD) affects ∼280 million people globally (1) and is a leading cause of disability (2). Approximately one third of patients do not respond to conventional monoaminergic antidepressants (3), highlighting the need for novel treatments targeting alternative mechanisms.

Accumulating evidence indicates that inflammation –defined as the activation of the innate immune system and release of pro-inflammatory proteins in response to insult or presence of a foreign pathogen– is implicated in the pathophysiology of MDD (4). Inflammatory markers include proteins such as C-reactive protein (CRP), interleukin (IL)-1, IL-6, and tumour necrosis factor (TNF)-α. Approximately one quarter of patients with depression show low-grade inflammation detected through elevated circulating inflammatory markers; an estimated 50% higher odds than matched healthy controls (5–7). Inflammatory abnormalities have also been detected in the central nervous system. Stress-induced peripheral inflammation can compromise the integrity of the blood-brain barrier (BBB), allowing inflammatory cytokines to enter the brain (8). Consistent with this, elevated cerebrospinal IL-6 and TNF-α, as well as elevated translocator protein binding, a neuroimaging marker of central inflammation, have been observed in patients with MDD compared to controls (9,10). Longitudinal observational studies have demonstrated that elevated IL-6 and CRP precede depressive symptoms, supporting a prospective association (11). Additionally, experimental induction of acute inflammation through proinflammatory agents including lipopolysaccharide (LPS) and interferon-α (IFNα) suggests that inflammation can contribute to depressive-like sickness behaviours in both humans and animals, including changes in sleep, fatigue, and anhedonia (12,13). Proinflammatory cytokines released in the context of immune activation may therefore contribute to the pathogenesis of MDD or a subset of depressive symptoms.

MDD is clinically heterogeneous and can be diagnosed through multiple symptom combinations (14). Its symptoms can be broadly classified into emotional (e.g., low mood, anhedonia, worthlessness, and suicidal ideation), neurovegetative (e.g., appetite gain/loss, sleep problems, fatigue), and cognitive (e.g., difficulty concentrating, psychomotor slowing) (15). In line with the sickness behaviour model of MDD, inflammation is most strongly associated with the neurovegetative and anhedonic symptoms of MDD (16), suggesting that different symptoms may have distinct aetiological pathways. Inflammation-related symptoms are clinically important, as anhedonia, fatigue, and cognitive problems are among the main contributors to functional impairment in MDD, alongside low mood (17). However, selective serotonin reuptake inhibitors (SSRIs) show limited efficacy for many of these symptoms, including anhedonia and neurovegetative symptoms (18–20), highlighting the need for new treatments tailored to these undertreated symptoms.

Clinical trials in chronic inflammatory conditions have reported beneficial effects for anti-cytokine treatments and non-steroidal anti-inflammatory drugs (NSAIDs) on mood (21,22). However, such studies focused on physical illness as the primary outcome, and improvements in patients’ physical condition may have mediated the observed benefits to mood. Some studies have examined the effectiveness of immunomodulatory medications for MDD as the primary outcome, with mixed results (23–26). Recent evidence indicates that anti-inflammatory approaches may specifically benefit treatment-resistant patients with elevated inflammation (27). Moreover, clinical trials tend to involve small samples (26) and the effectiveness of immunomodulatory medications may vary across MDD symptoms, potentially contributing to inconsistent findings (28). Beyond these mixed findings, many immunomodulatory drugs remain unexplored for MDD treatment. A systematic assessment and prioritisation of the effects of these compounds on MDD, including symptom-specific effects, is therefore needed.

Over 90% of existing drugs exert their effects by targeting proteins (29). Genetically informed methods, which leverage the fact that genes encode proteins, can therefore be used to identify proteins causally implicated in disease, including immune-relevant proteins (30). These proteins represent promising drug targets to prioritise the repurposing of existing immunomodulatory compounds. Mendelian Randomisation (MR) applied to drug repurposing can infer causality between immune-related drug targets and clinical outcomes. For *cis*-MR, genetic variants within or near the gene encoding a target protein of interest (*cis*-region) are selected as instruments to test whether the protein lies on the causal pathway to the disease (31). A causal relationship implies that pharmacological perturbation of the protein may be therapeutically effective (32). Two types of instruments are used; protein quantitative trait loci (pQTLs), which index protein values corresponding to the target (33), and expression quantitative trait loci (eQTLs) to infer the role of mRNA expression in the *cis*-region (34).

*Cis*-MR has previously identified potential MDD targets including NEU1, CD40, and IL-8 through studies of NSAIDs (35) and inflammation-related proteins (36). However, the full range of immunomodulating drugs, including those targeting innate and adaptive immune processes, host inflammatory pathways, and cytokine inhibitors, has not been examined. These compounds act on a broader range of immune pathways implicated in MDD beyond classical anti-inflammatory mechanisms. Anti-infectives, for example, primarily developed to bind bacterial targets, may also modulate host immune pathways. Moreover, prior *cis*-MR studies were restricted to plasma proteins, despite evidence of inflammation in brain and central nervous system tissue in MDD (9,10).

This study sought to evaluate immunomodulatory drug targets for evidence of an effect on MDD using *cis*-MR with blood and brain eQTL and pQTL datasets and the latest MDD GWAS (37). We subsequently prioritised targets and their corresponding compounds with the greatest repurposing potential based on the strength and consistency of genetic evidence and compounds’ expected therapeutic direction. Given symptom heterogeneity in MDD, we also tested whether targets were differentially associated with specific symptoms.

## Method

### Exposure & outcome data

#### Target selection

We selected approved immunomodulatory drugs based on Anatomic Therapeutic Codes (ATC) classification (denoting organ/system on which they act and their pharmacological properties), capturing compounds acting on immune and inflammatory pathways potentially implicated in MDD pathophysiology (4) (see Supplementary Methods 1). We identified 514 unique drugs, with target proteins for 204 of these identified using tractability data from ChEMBL (38,39), resulting in 123 unique targets selected for testing in *cis*-MR analyses (as multiple drugs can target the same protein). In ChEMBL, drugs targeting multi-subunit complexes may be annotated to a single target entry (39). However, Open Targets uses Ensembl gene identifiers as its primary target definition, such that each gene encoding a subunit is represented independently and constitutes a separate target in our analysis. The 310 drugs without targets for testing either have an unknown mechanism of action, or act solely on non-human targets. Gene ontology (GO) biological process annotations indicated that 39% of the testable targets are annotated to immune-related signalling processes, defined by the GO terms containing “immune”, “interleukin”, or “cytokine”.

#### Exposure and outcome

Genetic instruments for the exposure were derived from nine GWASs examining eQTLs or pQTLs in both blood (40–42) and multiple brain regions (Table 1) (43,44). We analysed each QTL dataset (or ‘panel’) independently to account for platform- and tissue-specific variation. To increase proteomic coverage, we included data from two different blood plasma pQTL GWAS, measured using different protein assay platforms (SomaScan and Olink) (40,41). We defined genetic instruments as variants located within 200kb of the gene encoding the target protein (45), with a minor allele frequency (MAF) >0.01, and associated with the genetic exposures at p < 1 × 10-6 (31). We removed variants with a stronger standardised effect on the outcome than on the exposure using MR Steiger filtering to limit potential reverse causation (46). Linkage disequilibrium (LD) pruning was then performed at an R2 threshold of 0.4, based on LD estimates from 5,000 UK Biobank participants of European genetic ancestry (47). A total of 122 targets had instruments that passed filtering criteria. For our primary outcome we used summary statistics from the PGC MDD phase 3 GWAS meta-analysis (excluding 23&Me, Ncases = 412,305, Ncontrols = 1,588,397) of European genetic ancestry (37). For the secondary outcomes, we used summary statistics from GWASs of eight individual depressive symptom items in the Patient Health Questionnaire-9 (PHQ-9) (Ns = 224,535-308,421) (48).

**Table 1.** pQTL and eQTL GWAS panels used for exposure instruments. Summary of the nine pQTL and eQTL GWAS datasets used to derive genetic instruments in the cis-MR analyses, including number of targets tested and counts of significant MR associations identified within each panel.

| QTL | Tissue type | Dataset (assay / meta-analysis) | N = | Number of unique significant drug targets / tested in MR |
| --- | --- | --- | --- | --- |
| pQTL | Blood plasma | deCODE (SomaScan) [PMID: 34857953] | 35,559 | 9/27 (33.3%) |
|  |  | UKB-PPP (Olink) [PMID: 37794186] | 54,219 | 6/18 (33.3%) |
|  | Brain (dorsolateral prefrontal cortex) | ROSMAP (Tandem Mass Tags) [PMID: 33571421] | 330 | 1/17 (5.9%) |
| eQTL | Whole-blood and peripheral blood mononuclear cell | eQTLGen (meta-analysis of 37 studies) [PMID: 34475573] | 31,684 | 31/98 (31.6%) |
|  | Brain/nervous system | MetaBrain (meta-analyses of up to 15 studies) [PMID: 36823318] | 208 | Basal ganglia: 13/78 (16.7%) |
|  |  |  | 492 | Cerebellum: 24/113 (21.2%) |
|  |  |  | 2683 | Cortex: 28/121 (23.1%) |
|  |  |  | 168 | Hippocampus: 5/64 (7.8%) |
|  |  |  | 285 | Spinal cord: 7/53 (13.2%) |

### Mendelian randomisation analysis

We conducted *cis*-MR analyses using a purpose-built Python package (49). For targets with a single genetic variant, effects were estimated using the Wald ratio estimator (MR-Wald). Where multiple genetic instruments were available, effect estimates were derived using a generalised least squares implementation of the inverse-variance weighted estimator (MR-IVW) (50) to account for residual LD. We applied heterogeneity-based pruning to exclude variants with large leverage (>3 times the mean) or outlier statistics (chi-square >11) to minimise potential bias from horizontal pleiotropy (49,51). We fit fixed-effects IVW models for targets with 3 instruments or less, otherwise fitting random effects (52,53). A false-discovery rate (FDR) correction was applied, accounting for 589 tests across the nine panels spanning brain and blood.

### Sensitivity analyses and target annotation

#### Robust MR

We further tested the significant IVW estimates from the primary analysis with three robust MR methods: contamination mixture (54), weighted median methods (55), and generalised least squares implementation of MR-Egger with heterogeneity-based pruning (56). We applied FDR correction for 285 tests. Robust MR analyses that yield significant effects in the same direction as IVW estimates provide additional support for the causal interpretation of the estimated effects (57).

#### Colocalisation

We performed colocalisation analyses for all targets that were significant in the main MR analysis to examine whether MDD and the molecular traits in question (pQTLs/eQTLs) are associated via shared or distinct causal variants. In the colocalisation framework, the posterior probability for hypothesis 3 (PPH3) indicates that mRNA/protein expression of the drug target and MDD have distinct causal variants, which leads to a violation of MR assumptions. Conversely, the posterior probability for hypothesis 4 (PPH4) indicates shared causal variants (45,58). A posterior probability of ≥0.8 was used as the threshold for both PPH3 and PPH4 to indicate robust evidence supporting each hypothesis (Supplementary Methods 2).

#### Estimated therapeutic relevance

For each compound acting on a given target, we inferred its expected therapeutic effect by combining the MR-estimated direction of effect with the compound’s pharmacological action data from ChEMBL (39). Compounds mechanistically countering the target’s risk-increasing effect on MDD (e.g. inhibitors of targets positively associated with MDD) were classified as having a beneficial therapeutic direction. Compounds mechanistically acting in the same direction as the target’s risk-increasing effect (e.g. inhibitors of targets negatively associated with MDD) were classified as having adverse effects. Compounds with mechanisms not specifying activation or inhibition were classified as “unknown”.

### Target prioritisation criteria

We prioritised targets with (i) consistent directional effects across multiple panels; (ii) IVW estimates supported by at least one significant robust MR estimate with concordant direction with the IVW estimate; and (iii) a compound with a predicted beneficial therapeutic effect in directionally consistent panels. For targets with significant MR estimates only in brain panels, we only prioritised these if their corresponding compounds cross the blood-brain barrier.

We deprioritised targets with directionally inconsistent significant effects between panels or robust estimators, or evidence for distinct causal variants in the colocalisation analysis (PPH3), and targets for which available compounds have unknown or adverse therapeutic effects.

### Individual symptoms analysis

We conducted secondary analyses to examine the associations between immunomodulatory drug targets and individual symptoms of MDD. We used the same exposure GWASs as the main *cis*-MR analysis. As outcomes, we used GWASs of eight individual depressive symptom items in the PHQ-9 (48). We excluded ‘psychomotor changes’ due to high genetic correlation with ‘cognitive problems’ (*r*_*g*_ = 0.98), choosing the latter for its greater number of genome-wide significant loci. Each symptom is rated on a 0-3 scale capturing past-fortnight frequency. To assess whether *cis*-MR effect estimates differed across depressive symptoms, we tested heterogeneity of symptom-level MR estimates using Cochran’s Q (59) using a multivariate random-effects framework. To account for phenotypic overlap between symptoms and shared genetic instruments, we conducted sensitivity analyses under alternative correlation assumptions (see Supplementary Methods 3). We assessed statistical significance using FDR correction.

## Results

### Primary MR analysis

Seventy-two unique drug targets (of 122 tested) were significantly associated with MDD in at least one of the nine exposure panels in the primary *cis*-MR analysis after FDR-correction (Supplementary Table 1). These correspond to 60 unique targets as defined by ChEMBL, where multi-subunit complexes are collapsed into single target entries, whereas our analysis considers each protein-encoding subunit separately. Hereafter, we refer to target components as putative targets. Among them, 20 met criteria for genetic robustness (robust MR evidence; no strong evidence for PPH3; directional consistency in ≥2 panels) (Figure 1). None of the targets had strong evidence for PPH4 (shared causal variants), although only six targets had sufficient power for colocalisation analyses. Thirteen of these are targeted by compounds predicted to have a beneficial therapeutic direction of effect for MDD (Table 2 and Supplementary Table 2). These 46 compounds, of which 28 are antihistamines targeting HRH1, are classified as high priority for repurposing for MDD (Supplementary Table 3).

**Table 2.** High priority drug targets and compounds with estimated beneficial therapeutic effect for MDD.

| Target (protein name) | Tissue (MR effect direction) | Significant symptoms (MR effect direction) | Estimated therapeutic MoA | Beneficial compounds | Primary indications | MDD-related effects |
| --- | --- | --- | --- | --- | --- | --- |
| C1S (complement component 1s subcomponent) | Basal ganglia eQTL (+), hippocampus eQTL (+), spinal cord eQTL (+), blood eQTL (+) | Anhedonia (+), negative thoughts (+), fatigue (+), sleep (+), cognitive problems (-) | Inhibition | Sutimlimab | Cold agglutinin disease | — |
| CRBN (protein cereblon) | Cortex eQTL (+), blood eQTL (+) | Anhedonia (+), cognitive problems (+), fatigue (+), sleep (+) | Inhibition | Pomalidomide<br>Lenalidomide (Cross BBB) | Cancer | Appetite, depression, fatigue, insomnia, mood, weight |
| CUL4A (Cullin-4a) | Cerebellum eQTL (+), cortex eQTL (+), blood eQTL (+) | Mood (-), cognitive problems (-), sleep (-), appetite (-) | Inhibition | Lenalidomide<br>Pomalidomide (Cross BBB) | Cancer | Appetite, depression, fatigue, insomnia, mood, weight |
| DPEP1 (dipeptidase 1) | Blood pQTLs (+) | Anhedonia (+), cognitive problems (+), fatigue (+), sleep (+), appetite (+)<br>Negative thoughts (-) | Inhibition | Cilastatin (Crosses BBB) | Bacterial infections | — |
| FCGRT (IgG receptor FcRn large subunit) | Cortex eQTL (+), blood eQTL (+) | Mood (+), anhedonia (+) | Inhibition | Efgartigimod alfa | Myasthenia gravis | — |
| FKBP1A (FKBP Prolyl Isomerase 1A) | Cerebellum eQTL (+), cortex eQTL (+) | Mood (+), negative thoughts (–), sleep (–) | Inhibition | Everolimus<br>Tacrolimus<br>Sirolimus<br>(Cross BBB) | Immunosuppression / organ transplant | Anxiety, appetite, depression, insomnia, mood, sleep, suicidal behaviours, weight |
| HRH1 (histamine H1 receptor) | Cortex eQTL (+), hippocampus eQTL (+) | Sleep (+)<br>Negative thoughts (–) | Inhibition | Antihistamines (see Supplementary Table 3 for full list)<br>(Cross BBB) | Allergic rhinitis | Anxiety, appetite, fatigue, insomnia, mood, restlessness, sleep, suicidal, weight |
| IL1RL2 (interleukin-1 receptor-like 2) | Blood pQTLs (+) | Sleep (+), mood (–), anhedonia (–), negative thoughts (–) | Inhibition | Spesolimab | Psoriasis (Fatigue) | – |
| IMPDH1 (Inosine-5'-monophosphate dehydrogenase 1) | Cortex eQTL, blood eQTL (+) | Mood (+), anhedonia (+), negative thoughts (+), fatigue (+) | Inhibition | Ribavirin | Hepatitis C | Anxiety, appetite, depression, mood, sexual, sleep, suicidal, weight |
| MMP7 (matrix metalloproteinase-7) | Blood pQTLs (+) | Mood (+), negative thoughts (+), suicidal ideation (+), cognitive problems (+), sleep (+) | Inhibition | Doxycycline<br>(Crosses BBB) | Acne vulgaris | Anxiety, appetite |
| POLE2 (DNA polymerase epsilon subunit 2) | Basal ganglia eQTL (+), cerebellum eQTL (+), cortex eQTL (+) | Mood (+), anhedonia (+), concentration (+), negative thoughts (+), cognitive problems (+), sleep, appetite (+) | Inhibition | Clofarabine<br>Cytarabine<br>Fludarabine<br>Gemcitabine<br>(Cross BBB) | Leukaemia | Anxiety, appetite, depression, fatigue, weight |
| PRIM1 (DNA primase small subunit) | Basal ganglia eQTL (+), cerebellum eQTL (+), blood eQTL (+) | Concentration (+), anhedonia (+) | Inhibition | Clofarabine<br>Cytarabine<br>Fludarabine<br>Gemcitabine<br>(Cross BBB) | Leukaemia | Anxiety, appetite, depression, fatigue, weight |
| S1PR4 (sphingosine 1-phosphate receptor 4) | Cerebellum eQTL (–), cortex eQTL (–) | Suicidal ideation (+), sleep (+), appetite (+), cognitive problems (–) | Activation | Fingolimod<br>(Crosses BBB) | Multiple sclerosis | Depression, sleep, weight |

*Note:* A positive MR estimate (+) indicates that higher target expression increases MDD risk, suggesting that beneficial compounds would have an inhibitory MoA on the target. A negative MR estimate (–) indicates that lower target expression increases MDD risk, suggesting that beneficial compounds would have an activating MoA. “Significant symptoms” refers to the symptoms for which the *cis*-MR estimate was significant in at least one exposure panel, along with the corresponding effect direction. “MDD-related effects” include MDD-related terms listed as side-effects of the compound, where multiple compounds are included, MDD-related effects are presented separately for each compound.
BBB, blood-brain barrier; eQTL, expression quantitative trait loci; MDD, major depressive disorder; MoA, mechanism of action; pQTL, protein quantitative trait loci.

**Figure 1.**
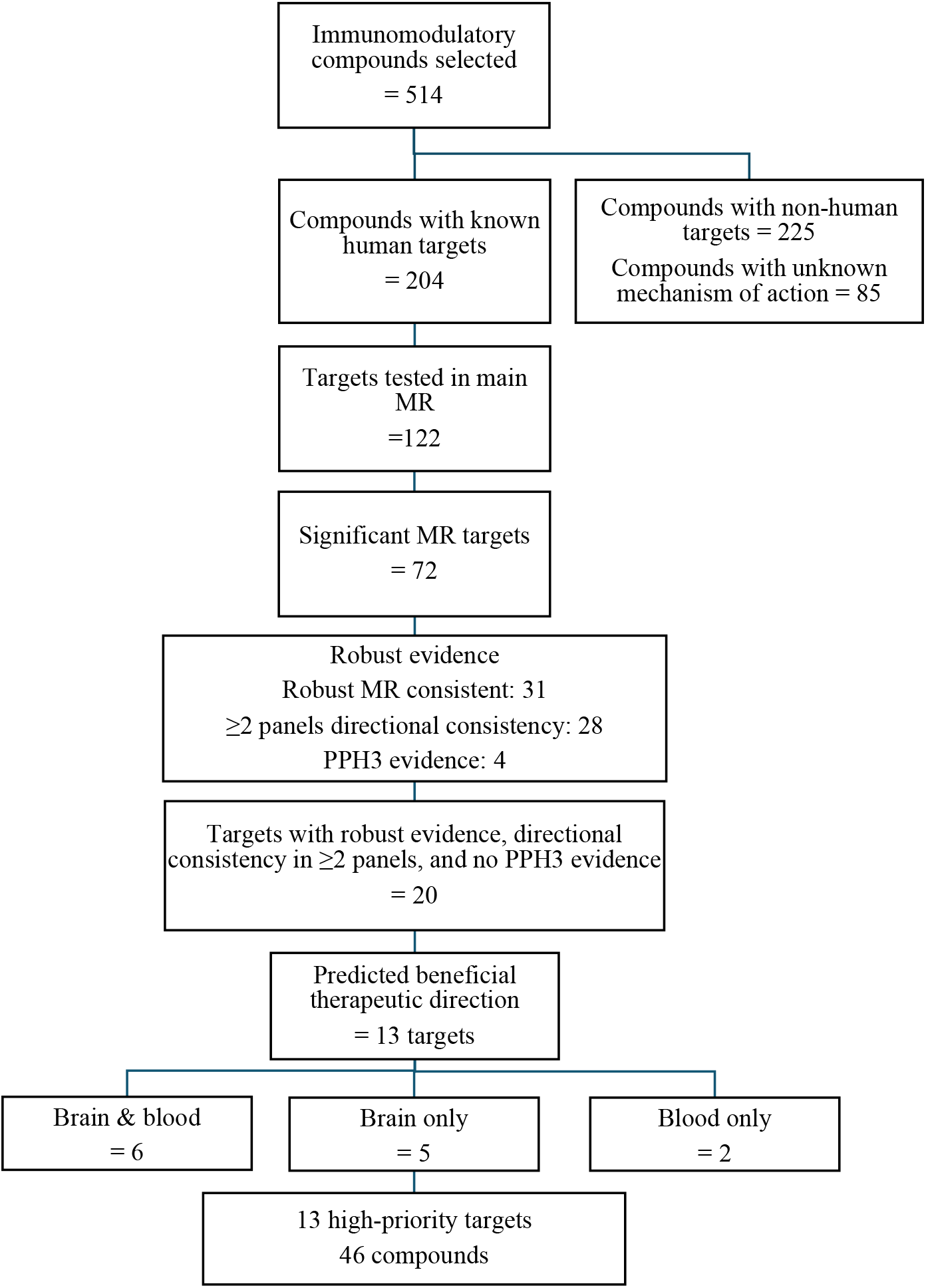
Target identification and prioritisation workflow. Flowchart showing identification of high-priority immunomodulatory compounds for repurposing for major depressive disorder (MDD)

### Individual symptoms

Of the 72 unique targets significant in the main MDD analysis, 60 were associated with at least one individual depressive symptom (Supplementary Table 1). The number of significant targets for each symptom ranged from 25 (suicidal ideation; appetite) to 34 (sleep problems). Five targets were significantly associated with all eight symptoms.

Figure 2 shows the estimated effects of each compound on individual symptoms, based on the panel in which the compound’s target showed the largest significant effect on MDD. Estimated effects by symptom in all panels are shown in Supplementary Figure 1. Under the primary symptom-correlation assumption (*r* = 0.5), 50 targets (83.3%) showed significant heterogeneity across symptoms. The number of heterogeneous targets varied depending on the assumed correlation structure. When no correlation was assumed (*r* = 0), 34 targets showed evidence of heterogeneity, whereas under a high correlation assumption (*r* = 0.8), 57 targets were heterogeneous. Among the prioritised targets, three (MMP7, IMPDH1 and POLE2) had significant effects in the same direction for more than half the symptoms. Four (CRBN, FCGRT, CUL4A, and PRIM1) had significant effects for up to four symptoms, and six (C1S, DPEP1, FKBP1A, HRH1, IL1RL2, and S1PR4) had effect estimates in opposite directions across symptoms, suggesting that modulation of these targets may exert both beneficial and adverse effects on different depressive symptoms.

**Figure 2.**
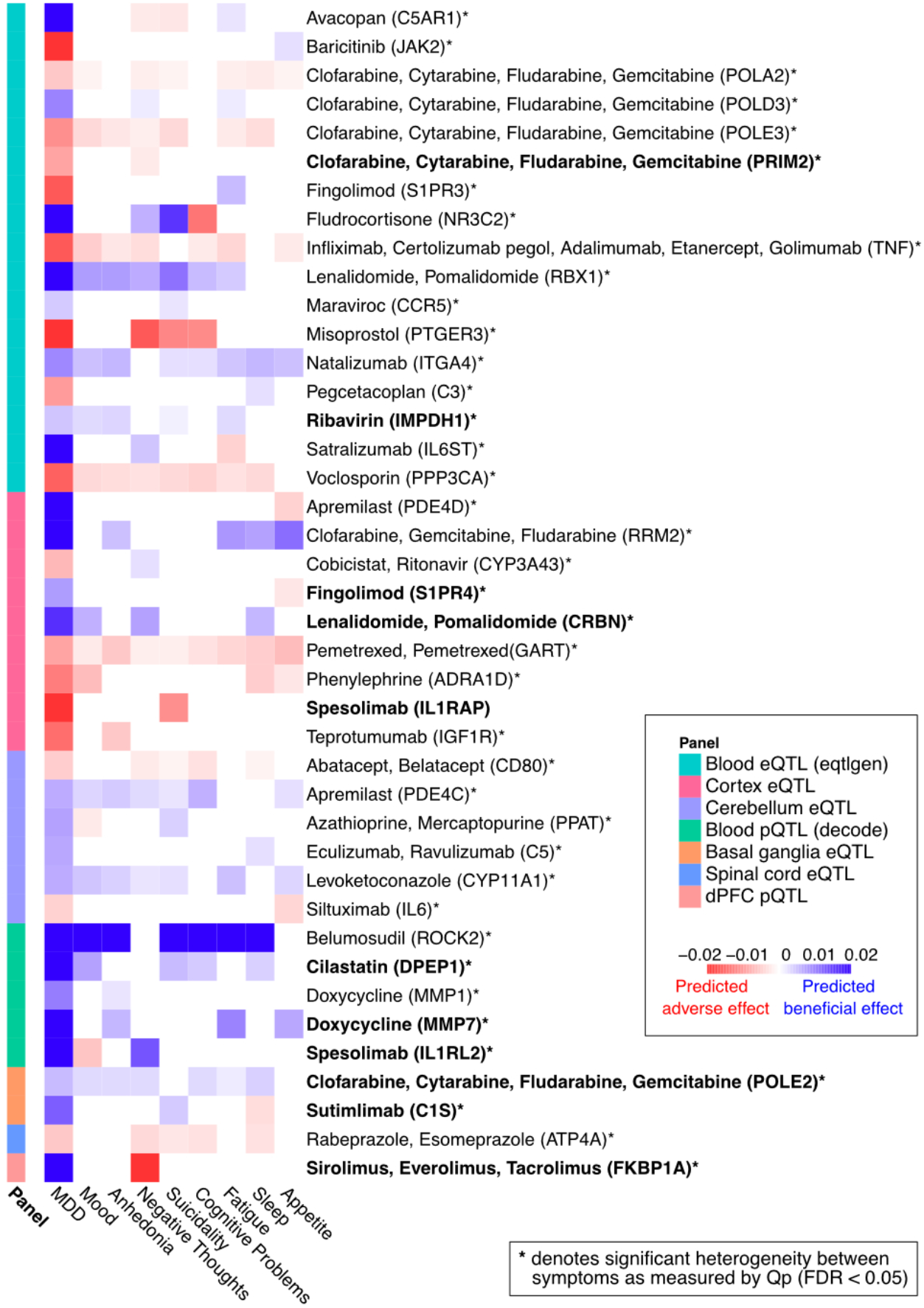
Predicted effects of immunomodulatory compounds on depressive symptoms. Heatmap showing the estimated effects of each compound on individual depressive symptoms, based on the *cis*-Mendelian randomisation effect of its target in the panel where it had the largest significant effect on major depressive disorder (MDD) diagnosis (case-control). Rows represent compounds (with the panel of strongest effect indicated), and columns represent depressive symptoms. Blue shades indicate predicted beneficial effects, red shades indicate predicted adverse effects, and white cells indicate non-significant associations. *Note:* bolded text indicates compounds with prioritised targets.

## Discussion

### High-priority compounds for MDD

We identified 46 high priority immunomodulatory compounds with targets significantly associated with MDD for potential repurposing. These compounds span drug classes targeting different immune pathways, including antibiotics, cytokine inhibitors, and immunosuppressants. Notably, over half these compounds were antihistamines, reflecting the large number of drugs targeting histaminergic signalling. Ten compounds, acting on targets not previously implicated in MDD, represent novel candidates for repurposing. The remaining compounds, or their targets, have been previously investigated for potential antidepressant effects, including doxycycline, fingolimod, and sutimlimab.

The tetracyclic antibiotic doxycycline targets matrix metalloprotease-7 (MMP7) has significant consistent effects for most symptoms, including mood, negative thoughts, and sleep problems. MMP7 is implicated in inflammation and BBB permeability (60,61), which may underlie doxycycline’s antidepressant-like effects in pre-clinical studies (62,63) and its risk-reducing effects for schizophrenia (64), although clinical studies examining doxycycline for MDD are lacking (65). Fingolimod, which modulates the prioritised target sphingosine-1-phosphate receptor (S1PR4), is used in multiple sclerosis (MS) and is also neuroprotective. Its beneficial effects on hippocampal neuron damage and BBB damage may be therapeutically beneficial for MDD (66–68). These effects are consistent with observed improvements in depressive symptoms in open-label studies with MS patients (69,70).

Two of the monoclonal antibodies we prioritised have prior evidence of relevance to MDD: sutimlimab, which inhibits C1S (complement component 1S), is used in cold agglutin disease (CAD), and spesolimab, an IL-1 receptor-like 2 (IL-1RL2) antagonist, is used in psoriasis. The former has been associated with improvements in mental health and fatigue in CAD patients (71), although its effects on MDD have not been examined in non-CAD populations. Our analysis suggests that alongside beneficial effects on anhedonia, negative thoughts, and fatigue, sutimlimab may worsen cognitive problems. The effects of spesolimab on mood have not been investigated, however it inhibits pro-inflammatory MAPK and NF-κB signalling pathways (72,73) that have been implicated in MDD (74,75). Notably, spesolimab is predicted to have beneficial effects on sleep problems but adverse effects on mood, anhedonia, and negative thoughts. While neither of these compounds is BBB permeable, their targets were associated with MDD in blood panels, indicating that peripheral perturbation may be beneficial.

The anti-cancer drugs pomalidomide and lenalidomide both target protein cereblon (CRBN) and cullin-4A (CUL4A), subunits of the same E3 ligase complex. Pomalidomide administration reduces TNFα levels and attenuates glutamate-mediated cytotoxic effects *in vivo* (76), inflammation-associated pathways that have been implicated in MDD (77,78). However, both compounds are also associated with significant adverse effects, including thromboembolism and hematologic toxicity (79,80), limiting their potential for direct repurposing for MDD. Nonetheless, evidence implicating their targets in MDD pathophysiology may inform the development of novel compounds with greater target selectivity and improved efficacy-tolerability profiles. While CRBN and CUL4A show concordant directions of effect, discordant effects among other subunits of the E3 ligase complex may indicate subunit-specific therapeutic potential,

Regarding antihistamines targeting the histamine H1 receptor (HRH1), preclinical evidence suggests that some first-generation compounds can modulate microglial activation, neuroinflammation, and depressive-like behaviours (81). However, HRH1 showed discordant symptom-level effects, which may reflect its predominantly neuromodulatory rather than immune cell-intrinsic mechanisms. In contrast, other prioritised targets act on slower, structural immune processes, including immune cell proliferation and survival, signal termination via the ubiquitin-proteasome system, and immune persistence.

Together, the prioritised targets identified converge on immune cell-intrinsic mechanisms governing proliferation, proteostasis, and immune persistence, supporting a model of MDD driven by sustained, poorly resolved inflammation, rather than classical cytokine excess.

### Symptom-level results

The largely heterogeneous target effects across symptoms align with evidence of distinct pathophysiology underlying different depressive symptoms (16,82). While some targets (28%) demonstrated directionally consistent effects across more than half the symptoms, others showed more symptom-specific effects. Five targets showed significant, directionally consistent effects across all targets, but were not prioritised: Three of these had evidence for PPH3, while two lacked robust MR evidence and/or replication across exposure panels. A subset of targets showed opposing effects across symptoms, indicating potential for adverse or symptom-worsening effects of drugs acting on these targets. We observed these opposing symptom-level effects within tissue panels and for both pQTL and eQTL instruments, suggesting that these patterns are unlikely to be only driven by differences between tissues or molecular proxies. Pharmacological modulation of these targets may be appropriate for individuals presenting with particular symptom clusters. Interestingly, immunomodulatory targets were not restricted to a single symptom category (e.g. neurovegetative), suggesting that inflammatory processes are implicated across symptom domains.

### Clinical implications

Our results highlight the importance of examining treatment effects at the symptom level, rather than solely relying on clinically and etiologically heterogenous MDD diagnoses. Recent clinical trials of anti-inflammatory drugs illustrate that using neurovegetative symptoms as primary outcomes (83) provides a more nuanced understanding of treatment efficacy. Overlooking symptom heterogeneity in treatment development risks potentially delivering treatments that are ineffective against the most impairing symptoms or that exacerbate certain symptoms of the disorder (84). A symptom-level approach aligns with the transdiagnostic framework of psychopathology (85) to identify interventions that may be beneficial across disorders and/or for specific symptom clusters. Notably, we predicted that several compounds examined here may have beneficial effects specifically on fatigue, a symptom that is prevalent, debilitating, and poorly addressed by current treatments (86,87).

### Strengths and limitations

Our approach enabled systematic evaluation of a large number of immunomodulatory drugs, moving beyond associative evidence to prioritise targets with potential causal relevance to MDD. Moreover, by including both blood and brain panels, this study captures both peripheral and central immune processes implicated in MDD, increasing relevance for pharmacological intervention. However, as per the *cis*-MR approach, the present study was limited to drugs for which the target is known. Additionally, 61% of the targets we investigated are not annotated to immune signalling processes based on GO biological process terms. Nonetheless, our method of target selection captures targets that may have downstream anti-inflammatory effects. A further limitation is the potential for effect flipping across tissues or cell types, as the direction of QTL effects can differ between tissues (88). When the disease-relevant tissue differs from the tissue in which the QTL was measured, the MR estimate may not reflect the true causal or therapeutically relevant direction of effect. This is particularly relevant for brain-mediated phenotypes such as MDD. However, unlike prior studies, we were not restricted to blood-derived instruments and incorporated large brain panels, although bulk tissue QTLs cannot resolve cell-type-specific effects. Finally, our findings demonstrate that symptom-level analyses provide novel insights relevant to drug development. However, our symptom-level analyses are less powered than for MDD because of the measure itself (single items referring to a short period, which introduces measurement error) and a smaller GWAS. Larger symptom-level GWAS, possibly including different measures of the same symptom are warranted.

### Conclusion

This study identified immunomodulatory compounds with strong genetic evidence supporting their repurposing for MDD, providing a basis for further preclinical and clinical evaluations. Variability in target effects across individual depressive symptoms highlights the importance of symptom-level analyses for guiding the development of more targeted therapeutic strategies.

## Acknowledgements

The authors acknowledge the use of the UCL Myriad High Performance Computing Facility (Myriad@UCL), and associated support services, in the completion of this work

## Ethical approval

The UK Biobank has ethical approval from the North West Multi-centre Research Ethics Committee as a Research Tissue Bank approval (REC reference: 21/NW/0157).

## Data availability

GWAS summary statistics for MDD are available from the Psychiatric Genomics Consortium at https://pgc.unc.edu/for-researchers/download-results/. GWAS summary statistics for PHQ-9 are available from https://zenodo.org/records/13828101. UK Biobank data are available through application at https://www.ukbiobank.ac.uk/enable-your-research/apply-for-access/. Open Targets tractability data can be downloaded from http://ftp.ebi.ac.uk/pub/databases/opentargets/platform/latest/input/target/tractability/. QTL datasets are available from: Blood plasma pQTL data: deCODE (https://www.decode.com/summarydata/), UKB-PPP (https://www.synapse.org/#!Synapse:syn51364943/). Brain pQTL data: ROSMAP (https://www.synapse.org/#!Synapse:syn24172458). Blood eQTL data: eQTLGen (https://www.eqtlgen.org/). Brain eQTL data: MetaBrain (https://www.metabrain.nl/).

## Author contributions

JJ: Conceptualisation, Methodology, Data curation, Formal Analysis, Software, Visualisation, Writing – Original Draft

ARTK: Conceptualisation, Methodology, Formal Analysis, Software, Supervision, Writing – Reviewing & Editing

JR: Writing – Reviewing & Editing

LAC: Methodology, Writing – Reviewing & Editing AP: Writing – Reviewing & Editing

SC: Software, Data curation, Writing – Reviewing & Editing

CF: Methodology, Software, Resources, Data curation, Writing – Reviewing & Editing

AFS: Methodology, Software, Resources, Data curation, Writing – Reviewing & Editing

JBP: Conceptualisation, Methodology, Supervision, Writing – Reviewing & Editing

