## Supplementary Tables for "Genetic evidence for repurposing immunomodulatory drugs for major depressive disorder"

### Supplementary Methods

1. Target selection

We included ATC categories containing the terms "anti-inflammatory", "anti-infective", "immune", "histamine", and "corticosteroid" to capture drugs acting on immune and inflammatory pathways potentially implicated in MDD pathophysiology. We excluded categories for topical or locally acting compounds and combination categories as they contain two or more active ingredients, precluding attribution of effects to a specific compound/target. The following ATC categories were included:

| ATC Organ System | ATC Category |
|---|---|
| Alimentary tract and metabolism | Antidiarrheals, intestinal anti-inflammatory/anti-infective agents / Intestinal anti-inflammatory agents (A07E) |
| Cardiovascular system | Lipid modifying agents / HMG-CoA reductase inhibitors (C10AA) |
| Dermatologicals | Antipruritics, incl. antihistamines, anaesthetics, etc. (D04) |
| Systemic hormonal preparations, excluding sex hormones and insulins | Corticosteroids for systematic use (H02) |
| Anti-infectives for systemic use | Antibacterials for systemic use (J01)<br>Antimycotics for systemic use (J04)<br>Antimycobacterials Antivirals for systemic use (J04)<br>Immunoglobulins (J06B) |

| | |
|---|---|
| Antineoplastic and immunomodulating agents | Antineoplastic agents / Antimetabolites (L01B)<br>Immunosuppressants (L04) |
| Musculo-skeletal system | Anti-inflammatory and antirheumatic products (M01A) |
| Nervous system | Analgesics / Antimigraine preparations / Corticosteroid derivatives (N02CB) |
| Respiratory system | Antihistamines for systemic use (R06) |
| Sensory organs | Ophthalmologicals / Anti-inflammatory agents (S01B)<br>Otologicals / Corticosteroids (S02B)<br>Ophthalmologicals and otologicals / Corticosteroids (S03B) |
| Various | Therapeutic radiopharmaceuticals / Anti-inflammatory agents (V10A) |

2. Colocalisation

We assessed the statistical power for the colocalisation analyses based on the sum of PPH3 and PPH4, with a power threshold of 0.8 needed for the analysis to be considered sufficiently powered (1). The analyses were conducted using purpose-built Python packages that incorporate the *coloc* R package (*coloc.abf* function) (2, 3).

3. Individual symptoms analysis

For each target and panel, we meta-analysed the eight symptom-specific MR estimates, accounting for non-independence due to phenotypic correlation between symptoms and shared genetic instruments. We performed sensitivity analyses under alternative assumptions about the correlation between symptom-level sampling errors ($r$ = 0, 0.5, 0.8). We examined heterogeneity using Cochran's Q (4), applying an FDR multiple testing correction within each sensitivity analysis (FDR-adjusted $p < 0.05$). The analyses were conducted using the *metafor* R package.

**Supplementary Figures**

Figure 1: **Predicted effects of immunomodulatory compounds on symptoms of depression in all exposure panels.** Heatmap showing the estimated effects of each compound on individual depressive symptoms, based on the *cis*-Mendelian randomisation effect of its target in all exposure panels. Rows represent compounds and columns represent depressive symptoms. Blue shades indicate predicted beneficial effects, red shades indicate predicted adverse effects, and white cells indicate non-significant associations.

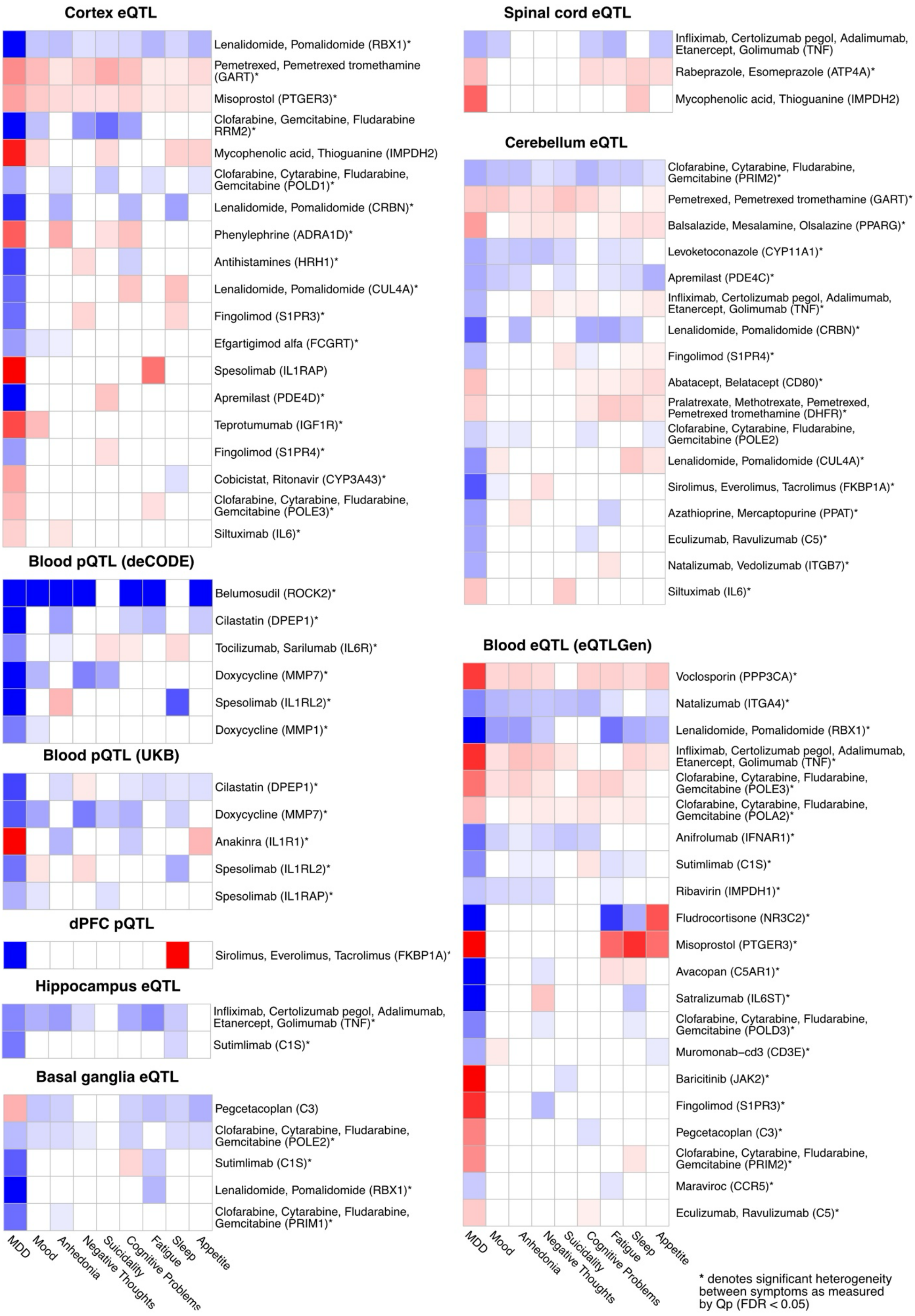
Cortex eQTL
Lenalidomide, Pomalidomide (RBX1)*
Pemetrexed, Pemetrexed tromethamine (GART)*
Misoprostol (PTGER3)*
Clofarabine, Gemcitabine, Fludarabine RRM2)*
Mycophenolic acid, Thioguanine (IMPDH2)
Clofarabine, Cytarabine, Fludarabine, Gemcitabine (POLD1)*
Lenalidomide, Pomalidomide (CRBN)*
Phenylephrine (ADRA1D)*
Antihistamines (HRH1)*
Lenalidomide, Pomalidomide (CUL4A)*
Fingolimod (S1PR3)*
Efgartigimod alfa (FCGRT)*
Spesolimab (IL1RAP)
Apremilast (PDE4D)*
Teprotumumab (IGF1R)*
Fingolimod (S1PR4)*
Cobicistat, Ritonavir (CYP3A43)*
Clofarabine, Cytarabine, Fludarabine, Gemcitabine (POLE3)*
Siltuximab (IL6)*
Blood pQTL (deCODE)
Belumosudil (ROCK2)*
Cilastatin (DPEP1)*
Tocilizumab, Sarilumab (IL6R)*
Doxycycline (MMP7)*
Spesolimab (IL1RL2)*
Doxycycline (MMP1)*
Blood pQTL (UKB)
Cilastatin (DPEP1)*
Doxycycline (MMP7)*
Anakinra (IL1R1)*
Spesolimab (IL1RL2)*
Spesolimab (IL1RAP)*
dPFC pQTL
Sirolimus, Everolimus, Tacrolimus (FKBP1A)*
Hippocampus eQTL
Infliximab, Certolizumab pegol, Adalimumab, Etanercept, Golimumab (TNF)*
Sutimlimab (C1S)*
Basal ganglia eQTL
Pegcetacoplan (C3)
Clofarabine, Cytarabine, Fludarabine, Gemcitabine (POLE2)*
Sutimlimab (C1S)*
Lenalidomide, Pomalidomide (RBX1)*
Clofarabine, Cytarabine, Fludarabine, Gemcitabine (PRIM1)*
MDD
Mood
Anhedonia
Negative Thoughts
Suicidality
Cognitive Problems
Fatigue
Sleep
Appetite
Spinal cord eQTL
Infliximab, Certolizumab pegol, Adalimumab, Etanercept, Golimumab (TNF)
Rabeprazole, Esomeprazole (ATP4A)*
Mycophenolic acid, Thioguanine (IMPDH2)
Cerebellum eQTL
Clofarabine, Cytarabine, Fludarabine, Gemcitabine (PRIM2)*
Pemetrexed, Pemetrexed tromethamine (GART)*
Balsalazide, Mesalamine, Olsalazine (PPARG)*
Levoketoconazole (CYP11A1)*
Apremilast (PDE4C)*
Infliximab, Certolizumab pegol, Adalimumab, Etanercept, Golimumab (TNF)*
Lenalidomide, Pomalidomide (CRBN)*
Fingolimod (S1PR4)*
Abatacept, Belatacept (CD80)*
Pralatrexate, Methotrexate, Pemetrexed, Pemetrexed tromethamine (DHFR)*
Clofarabine, Cytarabine, Fludarabine, Gemcitabine (POLE2)
Lenalidomide, Pomalidomide (CUL4A)*
Sirolimus, Everolimus, Tacrolimus (FKBP1A)*
Azathioprine, Mercaptopurine (PPAT)*
Eculizumab, Ravulizumab (C5)*
Natalizumab, Vedolizumab (ITGB7)*
Siltuximab (IL6)*
Blood eQTL (eQTLGen)
Voclosporin (PPP3CA)*
Natalizumab (ITGA4)*
Lenalidomide, Pomalidomide (RBX1)*
Infliximab, Certolizumab pegol, Adalimumab, Etanercept, Golimumab (TNF)*
Clofarabine, Cytarabine, Fludarabine, Gemcitabine (POLE3)*
Clofarabine, Cytarabine, Fludarabine, Gemcitabine (POLA2)*
Anifrolumab (IFNAR1)*
Sutimlimab (C1S)*
Ribavirin (IMPDH1)*
Fludrocortisone (NR3C2)*
Misoprostol (PTGER3)*
Avacopan (C5AR1)*
Satralizumab (IL6ST)*
Clofarabine, Cytarabine, Fludarabine, Gemcitabine (POLD3)*
Muromonab–cd3 (CD3E)*
Baricitinib (JAK2)*
Fingolimod (S1PR3)*
Pegcetacoplan (C3)*
Clofarabine, Cytarabine, Fludarabine, Gemcitabine (PRIM2)*
Maraviroc (CCR5)*
Eculizumab, Ravulizumab (C5)*
MDD
Mood
Anhedonia
Negative Thoughts
Suicidality
Cognitive Problems
Fatigue
Sleep
Appetite
* denotes significant heterogeneity between symptoms as measured by Qp (FDR < 0.05)
